# Rainfall and Temperature Modify Effects of On-Site Sanitation Intervention on *E. coli* Contamination in Bangladeshi Households

**DOI:** 10.1101/2024.08.22.24312444

**Authors:** Caitlin G. Niven, Mahfuza Islam, Anna Nguyen, Andrew Mertens, Amy J. Pickering, Laura H. Kwong, Mahfuja Alam, Debashis Sen, Sharmin Islam, Mahbubur Rahman, Leanne Unicomb, Alan E. Hubbard, Stephen P. Luby, Jessica A. Grembi, John M. Colford, Benjamin F. Arnold, Jade Benjamin-Chung, Ayse Ercumen

**Author notes:** Corresponding Author: Caitlin G. Niven, Jordan Hall Addition 2225, Raleigh, NC, 27606.

## Abstract

Weather events associated with climate change can influence the environmental spread and survival of fecal pathogens, potentially impacting the efficacy of water, sanitation, and hygiene (WASH) interventions. We used longitudinal data from a randomized controlled trial in Bangladesh to assess whether rainfall and temperature modified the effect of an on-site sanitation intervention on fecal contamination. Over 3.5 years, we enumerated *E. coli* in household samples along eight fecal-oral pathways (n=23,238 samples) and obtained daily weather data. The intervention included the provision of double-pit latrines, child potties, and scoops for removing child and animal feces, along with behavior change promotion. The intervention was associated with larger reductions in *E. coli* in/on mother hands, child hands, ponds, and flies (0.10-log to 0.91-log) following higher rainfall and in/on food, mother hands, child hands, soil, and ponds (0.11-log to 0.40-log) following higher temperatures, compared to drier and colder periods. The intervention slightly reduced *E. coli* in stored drinking water and had no consistent effect on *E. coli* in tubewell water, regardless of weather. Our findings suggest that sanitation interventions can help mitigate the effects of increased rainfall and temperature on environmental fecal contamination. Previous analyses of these data without stratification by daily weather only found small (approximately 0.10-log) reductions in *E. coli* in/on stored drinking water and child hands. Future WASH trials should incorporate weather data to identify periods of differential intervention effectiveness to understand how weather variability influences the outcomes of public health interventions and develop strategies to enhance resilience against climate change impacts in vulnerable communities.

**Research in Context:** *Evidence before this study:* Higher temperatures and increased rainfall linked to climate change have been associated with increased infectious disease transmission. Effective water, sanitation, and hygiene (WASH) interventions, such as access to improved sanitation facilities, are pivotal in reducing these risks. However, the efficacy of WASH interventions in interrupting fecal-oral transmission can be influenced by weather fluctuations, which can compromise service delivery and/or exacerbate environmental dissemination of fecal waste. We conducted a literature review on the impact of weather events and climate change on the environmental spread of fecal pathogens and their influence on WASH interventions. We used Google Scholar to identify articles published from 2000 to 2024 using the following search terms: (“climate change” OR weather OR weather events OR temperature OR heatwave OR rainfall OR precipitation) AND (sanitation OR “WASH intervention” OR “sanitation intervention” OR “intervention efficacy”) AND (pathogen OR enteropathogen OR “*Escherichia coli*” OR *“E. coli”* OR “fecal indicator” OR “fecal contamination” OR “fecal pathogens). Studies have documented the influence of weather conditions, particularly rainfall and temperature, on the effectiveness of WASH interventions in reducing fecal contamination and diarrhea. A recent meta-analysis found that lack of access to improved latrines and piped drinking water modified the association between rainfall/temperature and diarrhea incidence in children under the age of five but this analysis only focused on health outcomes and did not include measures of environmental contamination. In Bangladesh, WASH interventions, including water treatment and handwashing, were more effective in reducing *E. coli* contamination in stored drinking water and food during dry seasons compared to wet seasons, however, in a different longitudinal analysis focused on a sanitation intervention, seasonality did not significantly modify the relationship between the intervention and fecal contamination. However, these studies used seasonal definitions (wet vs. dry) which can miss transient weather fluctuations within a season. One study in Kenya used daily rainfall and temperature data to show that increases in drinking water contamination associated with increased rainfall were mitigated if households treated their water. No study has evaluated the influence of daily weather parameters on the effect of WASH improvements on a comprehensive set of fecal-oral pathways.

*Added value of this study:* This study utilizes longitudinal data from a randomized controlled trial that implemented water treatment, handwashing and sanitation interventions in Bangladesh (WASH Benefits), with over 23,000 *E. coli* measurements collected over 3.5 years. The measurements were taken along eight fecal-oral transmission pathways, including mother and child hands, stored food, stored drinking water, groundwater from tubewells, ponds, courtyard soil, and captured flies. Previous analyses of these data with no stratification by daily weather found only small reductions in environmental fecal contamination associated with the WASH interventions. In the present analysis, we re-analyzed these data using daily weather measurements to investigate whether rainfall and temperature modified the efficacy of the improved on-site sanitation intervention in reducing *E. coli* contamination in the domestic environment. Unlike previous analyses that did not consider daily weather variations, stratifying by daily weather conditions revealed differential intervention impacts on *E. coli* contamination, with larger reductions observed following periods of higher rainfall and temperature across multiple fecal-oral pathways (mother hands, child hands, food, soil, ponds, and flies).

*Implications of all the available evidence:* Understanding how interventions perform under varying weather conditions is crucial, particularly in low- and middle-income countries susceptible to extreme weather events. Integrating fine-grained weather data into sanitation intervention assessments can inform climate adaptation strategies. Our findings, combined with evidence from previous studies, indicate that on-site sanitation improvements, water treatment and broader WASH approaches can help protect against environmental fecal contamination associated with increased rainfall and temperatures. These insights are pivotal for designing resilient WASH interventions capable of mitigating climate-related health risks in vulnerable populations, supporting the Sustainable Development Goals in low- and middle-income countries. The WASH Benefits sanitation interventions implemented in rural Bangladesh demonstrated enhanced efficacy during periods of higher rainfall and temperature. These effects were not discernible in previous analyses that did not stratify by daily weather, where average reductions in *E. coli* counts were minimal, underscoring the importance of capturing weather variability in assessing intervention impacts. This approach also contrasts with earlier studies using calendar-based definitions of seasons, which may not fully capture the nuanced effects of climate variables on intervention efficacy. Collectively, these findings underscore the importance of incorporating fine-grained weather data in WASH trials to enhance the understanding of intervention efficacy under variable environmental conditions.

## Introduction

Higher temperatures and rainfall associated with climate change have been linked to increased risk of infectious disease, potentially through increasing the dissemination and survival of pathogens in environmental compartments, such as water and soil^1–4^. Water, sanitation and hygiene (WASH) interventions, such as improved access to clean water, sanitation facilities that isolate fecal waste from the environment, and adequate hygiene practices, are fundamental to preventing the spread of disease. However, the efficacy of these interventions can be influenced by environmental conditions, including weather fluctuations induced by climate change^4^. For example, heavy rainfall and/or flooding may impact service delivery or overwhelm sanitation systems, leading to increased fecal contamination of the environment, thereby undermining the effectiveness of sanitation interventions^5^. Alternatively, climate-resilient WASH interventions may deliver larger benefits during increased rainfall or temperature by protecting households from the effects of enhanced microbial dissemination and survival in the environment^3,6^. Resilient interventions that remain effective under weather-related stressors can ensure sustained access to clean water and effective sanitation in the face of climate change. In low- and middle-income countries vulnerable to extreme weather events, it is important to understand how the efficacy of WASH interventions may vary under different weather conditions^7^.

Here, we use longitudinal data from a randomized controlled trial in rural Bangladesh (WASH Benefits) that implemented individual and combined WASH interventions to assess the effect of rainfall and temperature on intervention effectiveness against fecal contamination in the environment. The interventions reduced diarrhea and enteric infections among young children^8,9^. However, a longitudinal analysis focused on the sanitation intervention demonstrated that the health benefits were confined to the rainy seasons; the intervention did not affect child diarrhea during dry seasons^10^. Similarly, a recent analysis of the trial data using daily rainfall and temperature values found that the WASH interventions reduced diarrhea prevalence by 51% following periods with heavy rainfall, compared to 13% following periods without heavy rainfall, and by 40% following periods with above-median temperature, compared to 9% following periods with below-median temperature^11^.

Investigating the underlying environmental routes of fecal contamination under different weather conditions is important for interpreting these health effects and understanding how intervention effectiveness can be improved under adverse weather conditions. Previous evaluations of the effects of the WASH Benefits interventions on fecal contamination found that the water treatment, handwashing and combined WASH interventions reduced *E. coli* in stored drinking water and food, with evidence of greater reductions during the dry season^12,13^. The sanitation intervention did not reduce *E. coli* in the domestic environment when measured approximately 4 months after intervention initiation^12^. In a longitudinal study conducted between 1 and 3.5 years after intervention initiation, the sanitation intervention slightly reduced *E. coli* counts in stored drinking water and child hand rinses, with no effect modification by season^14^. However, two of these studies defined seasons based on calendar month, and one study defined the monsoon season based on spatiotemporal rainfall data. It is important to consider granular rainfall and temperature measurements rather than general/binary seasonality as this approach can capture discrete weather events as well as intra-annual variation in the onset and duration of dry and wet seasons, which are expected to shift with climate change.

This current study re-analyses data from environmental samples collected over 3.5 years in the sanitation and control arms of the WASH Benefit Bangladesh trial using daily weather data to investigate whether rainfall and temperature modify the effectiveness of the sanitation intervention on fecal contamination in the domestic environment. Understanding how weather patterns modify the efficacy of WASH interventions is essential for strengthening resilience, long-term sustainability, and adaptive capacity in vulnerable communities as well as inform intervention designs for WASH under climate change^15^.

## Methods

### Study Design and Population

The WASH Benefits randomized controlled trial in rural Bangladesh (NCT01590095) aimed to assess the impact of water, sanitation, hygiene, and nutrition interventions on child health outcomes. The trial enrolled 5,551 pregnant women and followed their birth cohort (index children) for two successive years. Six to eight enrolled households formed a cluster, and eight adjacent clusters formed a block. Within each block, clusters were randomly assigned to intervention or control arms. The current study is focused on the sanitation intervention, which consisted of installing new or upgraded double-pit latrines with concrete-lined pits, concrete slabs and water seals, providing sani-scoops to remove animal and child feces from the compound, and plastic potties for children that were too young to use the latrine. The hardware was accompanied by regular visits by trained community health promoters to intervention households to promote improved sanitation practices. Intervention uptake was high and sustained^16,17^. Details of the study design, enrollment criteria, interventions and uptake assessment have been previously published^18^.

### Environmental Data Collection

Environmental samples were collected from households in the control and sanitation intervention arms over nine sampling rounds spanning 3.5 years. Sampling pathways included index child and mother hand rinses, stored food for children under five years old (primarily rice), stored drinking water, source water (from tubewells), ponds, courtyard soil, and captured flies. Samples were processed at the field laboratory of the International Centre for Diarrheal Disease Research, Bangladesh (icddr,b) within 24 hours of collection. The most probable number (MPN) of *E. coli* was enumerated using IDEXX Quanti-Tray/2000 with Colilert-18 media and incubation at 45°C^19^. Sample collection and processing details have been previously reported^12,14,20^.

### Weather Data

Daily rainfall observations were obtained from the GloH2O’s Multi-Source Weighted-Ensemble Precipitation (MSWEP) dataset, and daily temperature observations were obtained from the National Aeronautics and Space Administration’s Famine and Early Warning Systems Network (FEWS NET) Land Data Assimilation System for Central Asia (FLDAS-Central Asia) dataset. These datasets had resolutions of 1.0° updated every 3 hours and 0.01° updated daily, respectively. Dates with missing weather observations (0.3% (2/652) of study days) were imputed using available data for the same date from nearest neighboring household in the study^21^. Variables to classify rainfall and temperature were generated for antecedent periods of two and seven days before sample collection. Rainfall and temperature observations were classified as ‘extreme’ if any day in the antecedent period exceeded the 90th percentile of all daily observations occurring during the 3.5-year study period in the study area. Additionally, as a sensitivity analysis for this threshold, we classified observations as ‘heavy rain/elevated temperature’ if any day in the antecedent period exceeded the 80th percentile of daily rainfall and temperature values, respectively. We further evaluated whether rolling average rainfall and temperature during the antecedent periods was above the median (≥50th percentile of rolling average values) because the classifications based on the 80^th^ and 90^th^ percentiles of rainfall/temperature can lead to data sparsity when further stratified by study arm and sample type.

### Statistical Analysis

We estimated differences in log10-transformed *E. coli* counts between the sanitation intervention and control arms using generalized linear models (GLM) with a Gaussian distribution and robust standard errors to account for clustering by study block and repeated observations. Non-detects were imputed as half of the lower detection limit and values above the upper detection limit were imputed as the upper detection limit; detection limits varied by sample type due to differences in processed sample amounts and dilutions^12^. Models included an indicator variable for study block to account for geographical matching. Our analyses did not adjust for additional variables because random assignment balanced potential confounders between intervention and control arms^9,14^. We assessed effect modification by the weather variables above by including an interaction term between the study arm and each binary weather variable. We compared the magnitude of intervention effects across the different weather strata and interpreted p-values <0.20 for the interaction variable as evidence of significant effect modification^22^. Analyses were conducted in R (version 4.3.1, RStudio 2023.06.0+421).

## Results

Between July 2013 and December 2016, we collected 5740 stored drinking water, 1629 stored food, 5397 mother hand rinse, 6494 child hand rinse, 1098 source water (tubewell), 1928 soil, 557 pond, and 395 fly samples from 1243 unique households in the control and sanitation arms of the WASH Benefits Bangladesh trial (Table S1). The 90^th^ percentile of daily rainfall (classified as extreme rainfall) corresponded to 28.20 mm and the 80^th^ percentile (classified as heavy rainfall) corresponded to 16.44 mm. The median of rolling average rainfall values for two- and seven-day periods were 0.27 mm and 1.10 mm, respectively. The 90^th^ percentile of daily mean temperature (classified as extreme temperature) corresponded to 30.21°C and the 80^th^ percentile (classified as elevated temperature) corresponded to 29.29°C. The median of rolling daily average temperature values for two- and seven-day periods were 27.46°C and 27.52°C, respectively. The highest rainfall occurred between the months of April–October and the highest temperatures occurred between the months of March–September (Fig S1, Fig S2). Of 23,238 total samples, 12.0% (2779) were collected within two days and 23% (5342) within seven days after extreme rainfall, and 14.1% (3282) were collected within two days and 20.5% (4765) within seven days after extreme temperature (Table S2).

Across all observations (without stratification by weather), the sanitation intervention was associated with an approximately a 0.1-log reduction in *E. coli* counts in stored drinking water (Δlog10 = −0.08 (−0.14, −0.01), p-value= 0.03) and on child hands (Δlog10 = −0.06 (−0.13, 0.00), p-value=0.05) (Table 1). The intervention did not significantly reduce *E. coli* contamination in food, mother hands, soil, tubewell water, ponds, and flies.

**Table 1.** Intervention effects from improved on-site sanitation on log10-transfomed *E. coli* counts by sample type, using all study observations without stratification by weather.

|  | **Δlog10 (95% CI)** | **P-value** |
| --- | --- | --- |
| **Stored Water** | **-0.08 (-0.14, -0.01)** | **0.03** |
| **Food** | 0.06 (-0.08, 0.21) | 0.39 |
| **Mother Hands** | -0.02 (-0.10, 0.05) | 0.51 |
| **Child Hands** | **-0.06 (-0.13, 0.00)** | **0.05** |
| **Soil** | -0.06 (-0.17, 0.05) | 0.28 |
| **Source Water** | -0.05 (-0.13, 0.03) | 0.20 |
| **Ponds** | 0.08 (-0.21, 0.05) | 0.24 |
| **Flies** | -0.20 (-0.52, 0.12) | 0.22 |

Overall, rainfall modified the effect of the sanitation intervention on *E. coli* counts on mother hands, on child hands, in ponds, and in/on flies; the intervention consistently led to larger reductions in *E. coli* in these sample types following periods of higher rainfall. When extreme rainfall occurred within seven days prior to sampling, the intervention was associated with a 1-log reduction in *E. coli* in/on flies (Δlog10 = −0.91 (−1.65, −0.17)); there was no intervention effect in the absence of extreme rainfall (interaction p-value=0.03) (Fig 1, Table S3). Trends for flies were similar for extreme rain occurring within two days prior to sampling (with borderline significant interaction p-value), heavy rain occurring within two or seven days prior to sampling, and above-median rolling rainfall within two or seven days prior to sampling (interaction p-values<0.20) (Figs 1 and 2, Fig S3, Tables S3-S5). Extreme or heavy rainfall did not modify intervention effects on any other sample type during either antecedent period (interaction p-values >0.20) (Figs 1 and Tables S3 and S5). When the rolling average rainfall within two or seven days prior to sampling was above median, the intervention was associated with an approximately 0.1-log reduction in *E. coli* on child hands while there was no intervention effect when rolling average rainfall was below median, with a similar trend for *E. coli* on mother hands (interaction p-values<0.20) (Fig 2, Table S4). When the rolling average rainfall within seven days prior to sampling was above median, the intervention was also associated with a 0.25-log reduction in *E. coli* in ponds (Δlog10 = −0.25 (−0.48, −0.03)), with no effect when rolling average rainfall was below median (interaction p-value=0.10) (Fig 2, Table S4). Trends for ponds were qualitatively similar for all rainfall classifications and antecedent periods, with intervention effects appearing to be larger in magnitude following higher rainfall (interaction p-values ranging from 0.10 to 0.45). Rainfall of any classification over either antecedent period did not modify intervention effects on *E. coli* in stored drinking water, food, soil and tubewell water.

**Figure 1.**
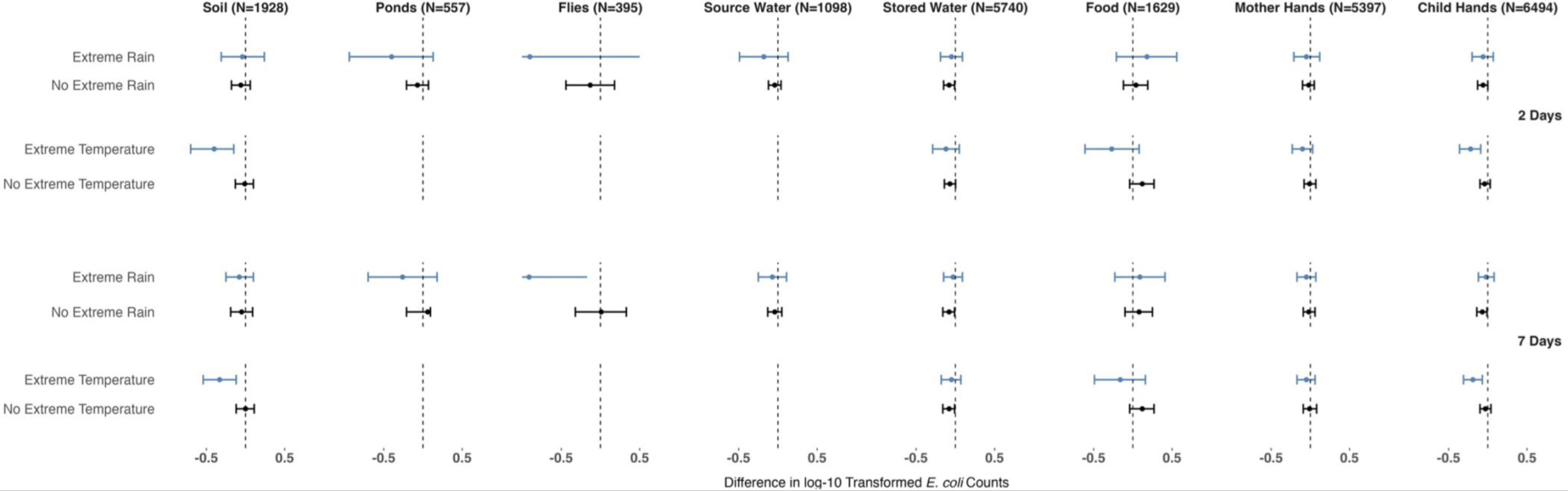
Forest plot of intervention effects from improved on-site sanitation on log10-transformed *E. coli* counts by sample type in strata of extreme rainfall and temperature ^a^ during 2- and 7-day antecedent periods ^b^. ^a^ Extreme rainfall defined as ≥ 28.20 mm, extreme temperature defined as ≥ 30.21°C. We could not estimate intervention effects on source water (tubewells), ponds and flies following periods of extreme temperature due to data sparsity. ^b^ Confidence intervals truncated for 2- and 7-day periods of extreme rainfall for flies, Δlog10 transformed *E. coli* counts and 95% CI: −0.90 (−2.30, 0.50) and −0.91 (−1.65, −0.17), respectively.

**Figure 2.**
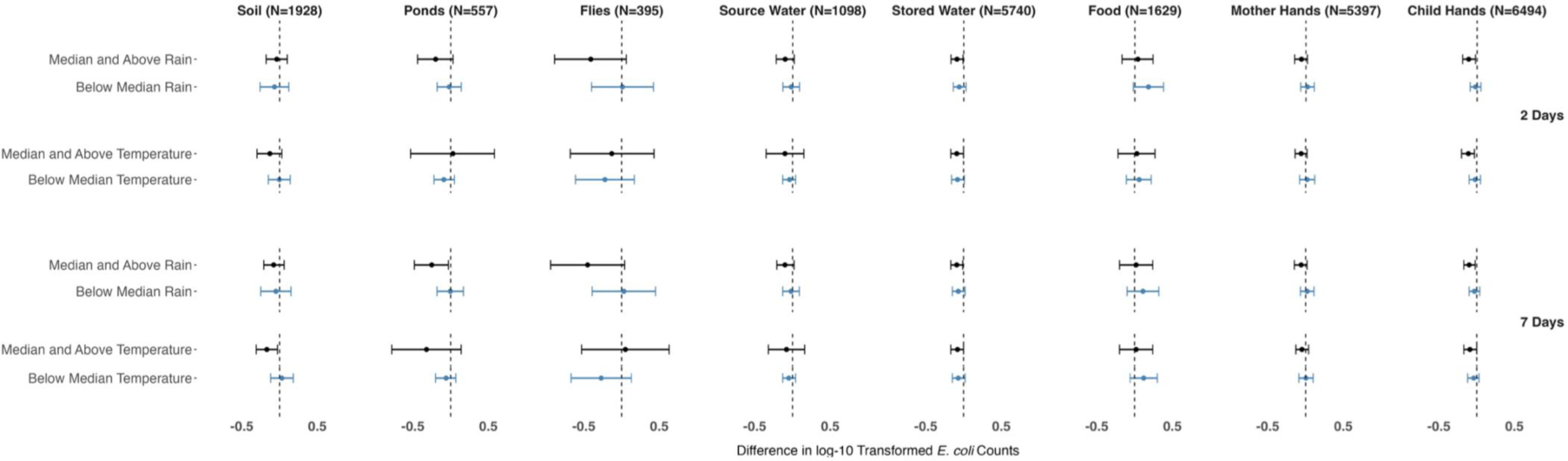
Forest plot of intervention effects from improved on-site sanitation on log10-transformed *E. coli* counts by sample type in strata of above and below median rolling average rainfall and temperature ^a^ during 2- and 7-day antecedent periods. ^a^ Median 2-day rolling average rain defined as ≥ 0.27 mm, median 2-day rolling average temperature defined as ≥ 27.46°C, median 7-day rolling average rain defined as ≥ 1.10 mm and median 7-day rolling average temperature defined as ≥27.52°C.

Overall, temperature modified the effect of the sanitation intervention on *E. coli* counts in food, on mother hands, on child hands, in soil, and in ponds, with the intervention consistently leading to larger reductions in *E. coli* in these sample types following periods of higher temperature. When extreme temperature occurred within two days prior to sampling, the intervention was associated with a 0.22-log reduction in *E. coli* on child hands (Δlog10 = −0.22 (−0.36, −0.09), interaction p-value=0.01) and 0.40-log reduction in *E. coli* in soil (Δlog10 = −0.40 (−0.70, −0.15), interaction p-value=0.01) (Fig 1, Table S3). For *E. coli* in food and on mother hands, there were no significant intervention effects in either temperature stratum, but the point estimates indicated larger reductions when extreme temperature occurred within two days prior to sampling (interaction p-value=0.05 for food, 0.18 for mother hands) (Fig 1, Table S3). These trends were similar for extreme temperature during the seven-day antecedent period (except for mother hands, Fig 1, Table S3) and for elevated temperature during either antecedent period (Fig S3, Table S5). Additionally, when elevated temperature occurred within two days prior to sampling, the intervention was associated with a 0.40-log reduction in *E. coli* counts in ponds (Δlog10 = −0.38 (−0.59, −0.17), interaction p-value=0.01), with similar findings for the seven-day period (Fig S3, Table S5). When the rolling average temperature was above median within two or seven days prior to sampling, the sanitation intervention was associated with larger reductions in *E. coli* counts on mother hands (interaction p-value=0.17), child hands (interaction p-value=0.05) and soil (interaction p-value=0.05) (Fig 2, Table S4). No measure of temperature modified intervention effects on *E. coli* in stored drinking water, tubewell water, or flies.

## Discussion

The WASH Benefits sanitation intervention, including double-pit latrines, child potties and child and animal feces management tools, reduced fecal contamination in rural Bangladeshi households more effectively following periods of higher rainfall and temperature. Across different classifications and antecedent periods, rain modified intervention effects on *E. coli* counts for four out of the eight sample types (mother and child hands, ponds, and flies) while temperature modified intervention effects on *E. coli* counts for five out of the eight sample types (food, mother and child hands, soil, and ponds). Reductions in *E. coli* counts ranged from 0.10 log on child hands to 0.91 log in flies following increased rainfall, and from 0.11 log on child hands to 0.40 log in soil following increased temperature. In contrast, analyses without stratifying by daily weather found small (<0.10 log) reductions in *E. coli* counts only for stored drinking water and child hands associated with the intervention, indicating that averaging across the study period concealed intervention effects taking place following increased rainfall and temperature. These findings can shed light on the mechanism behind the larger reductions in child diarrhea observed in the parent trial during the rainy seasons and following episodes of higher rainfall and temperature^23^. A re-analysis of diarrhea outcomes from the parent trial also found that the largest health benefits occurred among the lowest socioeconomic strata during the monsoon season, indicating joint effects from socioeconomic position and vulnerability to climate-sensitive pathogen transmission^24^. Our findings are consistent with a recent meta-analysis that found that when households lack access to improved latrines and piped drinking water, increasing temperature and rainfall are associated with increased incidence of diarrhea in children under the age of five^4^.

Higher rainfall can lead to increased initial dissemination (e.g. overflowing latrine pits) and subsequent dilution of fecal contamination in the environment^25,26^; these patterns, in return, can influence how effectively a sanitation intervention contains fecal contamination. For example, in our previous analysis, we found that increased rainfall was associated with increased *E. coli* contamination in ponds but decreased contamination in tubewell water and courtyard soil^21^. These patterns may indicate that, during heavy rain, latrine pits overflow into the surroundings and/or leak into the subsurface but fecal waste is then flushed out of the soil matrix into ponds with run-off ^27^. The larger reduction from the sanitation intervention in pond *E. coli* counts following increased rainfall in the current analysis suggests that the sanitation upgrades may have reduced rainfall-associated release of fecal waste from latrines into ponds. Possible mechanisms include that the concrete lining of the improved pits reduced leakage or that the double-pit design prevented overly full pits that are more likely to overflow with rain. Larger reductions from the sanitation intervention in *E. coli* counts in/on flies and on mother and child hands following increased rainfall also support the possibility that the intervention reduced rainfall-associated dissemination of fecal waste from latrines into the environment and subsequent vector/hand contact with fecal contamination.

Our previous findings also indicated increased contamination of stored drinking water and food following extreme rainfall^21^. However, the sanitation intervention was not any more or less effective against contamination of these pathways following increased rainfall. These findings suggest that the mechanisms for rainfall-associated contamination of stored drinking water and food (e.g. handling and storage conditions) were not influenced by sanitation improvements, despite the observed reductions in mother and child hand contamination after increased rain, which would be expected to reduce contamination of both stored water and food^28^. Drinking water treatment and safe storage of water and food following increased rainfall may protect against rainfall-associated contamination. A recent study in Kenya found that household water treatment mitigated the effects of rainfall on drinking water quality^29^.

Warmer temperatures can stimulate microbiological activity, potentially creating ideal incubation environments for vectors and fecal pathogens, which could be mitigated by sanitation interventions that isolate feces from the environment^3,30^. Conversely, warmer temperatures can also be associated with more sun exposure, which can inactivate microorganisms^3,6^. Our previous analysis found that higher temperatures were associated with increased contamination of stored drinking water and food in this setting^21^. In our current analysis, the sanitation intervention appeared to more effectively reduce food contamination following higher temperatures, but the reductions could not be distinguished from the null in any temperature stratum. There was a similar pattern for mother hands. The sanitation intervention was associated with a larger reduction in child hand contamination following increased temperatures, as well as larger reductions in contamination of soil and ponds. These findings may indicate that improved containment of fecal waste can differentially reduce proliferation of fecal organisms in the environment during warmer conditions, which may translate to less contaminated child hands.

It is also possible that rainfall and temperature affect adherence to sanitation practices, including the use of latrines, location of defecation, and proper disposal of child and animal feces^31^. Understanding the user convenience of sanitation hardware and any behavior changes during different weather conditions will be helpful to improve the implementation and adoption of sanitation practices and intervention strategies^32^. During increased rainfall or higher temperatures, individuals may change their latrine usage behavior, due to damage or resource conservation, or modify their defecation practices to avoid discomfort or inconvenience, potentially affecting levels of fecal contamination in the environment^26,33^. For example, during heavy rain, young children may be more likely to defecate in a potty indoors than openly defecate outdoors and similarly, older children and adults may be more likely to use latrines than open defecate. Domestic animals might be confined to different spaces during heavy rain and/or higher temperatures, thereby impacting where their feces are disposed of and subsequent levels of environmental contamination.

Our findings differ from previous assessments of the WASH Benefits interventions, which found that the water treatment, handwashing and combined WASH interventions more effectively reduced *E. coli* in stored water and food during the dry season compared to the wet season^12,13^ and no effect modification by season in sanitation intervention effects on environmental contamination^14^. The calendar-based (June-October) and/or binary definitions of wet season in these studies do not capture rain-free episodes during the wet season or occurrences of rain during the dry season. They also do not capture temperature fluctuations that occur within the same season and cannot differentiate between joint variations in rain and temperature (e.g., generally warmer temperatures coinciding with the wet season in Bangladesh). These factors can potentially explain the different findings from our current analysis that used daily weather data. As weather patterns shift under climate change and longstanding definitions of season no longer hold empirically, using finer grained weather data can provide improved insights over relying on season definitions. However, in regions like Bangladesh, where a distinct monsoon season can be reliably identified using rainfall data, using seasonality as an effect modifier could still be valuable^24^. This approach can distill a complex set of environmental/temporal factors into a single variable definition and help capture the combined influence of rainfall and temperature which might be missed if each factor is modelled separately. Operationally defining a monsoon season can also simplify program implementation and messaging (e.g. deliver and emphasize interventions only or more heavily during the monsoon season).

Our findings also offer nuance to previously published findings that only indicated small reductions in stored drinking water and child hand contamination from the sanitation intervention but no other effects on the broader domestic environment. Our current analysis indicates additional significant sanitation intervention effects – including reductions in *E. coli* contamination of ponds, courtyard soil, and flies – that are confined to periods of increased rainfall and/or temperature. Future research should use fine-grained and location-specific rainfall and temperature data to quantify the efficacy of sanitation and WASH interventions during different weather conditions to detect effects that may be concealed when averaged over the study period^29^. Longitudinal assessments spanning multiple seasons and a range of weather conditions can provide comprehensive insights into intervention effectiveness and inform strategies for maximizing impact under fluctuating weather conditions. Future WASH intervention studies should also consider targeted environmental sampling following periods of heavy rain and elevated temperatures. However, we also note that even when the sanitation intervention resulted in significant reductions in contamination under some weather conditions, *E. coli* counts in the environment remained relatively high among intervention recipients^12^.

This study leveraged an extensive longitudinal dataset to understand how daily weather variability affects the impact of multi-faceted sanitation improvements on several fecal-oral transmission pathways. However, the study also has limitations. We could not assess effect modification by extreme temperature for source water (tubewells), ponds, and flies because only a small number of samples coincided with periods of extreme temperature for these sample types. However, our sensitivity analysis using elevated instead of extreme temperatures supported our primary conclusions. Additionally, generalizing our findings to other settings may be limited by variations in environmental factors, socioeconomics, infrastructure, and cultural practices. While this study focused on rural households in Bangladesh, the specific underlying mechanisms driving intervention effects during different rainfall/temperature patterns may not apply to other settings.

Our findings indicate increased protection from sanitation improvements following periods of increased rainfall and temperature, suggesting that sanitation and potentially other WASH approaches can support efforts to mitigate adverse health impacts from climate change. This study contributes to the growing body of literature on climate change adaptation and incorporating climate concerns in the context of public health, providing insights for policymakers and practitioners to design context-specific sanitation interventions that remain effective in the face of evolving climatic conditions. Our findings hold significant implications for policy and public health practice, particularly in low- and middle-income countries facing climate-related health risks. Adaptive management strategies capable of addressing changing environmental conditions and robust interventions that are resilient to weather fluctuations are essential to mitigate impacts from climate variability. By integrating climate resilience into sanitation policy and practice, governments and organizations can better safeguard public health and foster sustainable development in vulnerable communities.

## Data Availability

All data produced are available online at https://osf.io/6u7cn.

