## Supplementary Materials for "Rainfall and Temperature Modify Effects of On-Site Sanitation Intervention on *E. coli* Contamination in Bangladeshi Households"

### Table of Contents

***Table S1:*** Number of samples collected by sample type and study round ______ **Page 2**

***Table S2***: Percentages of samples within each weather category across different antecedent timeframes ______ **Page 2**

***Table S3***: Intervention effects from improved on-site sanitation on log10-transformed *E. coli* counts by sample type in strata of extreme rainfall and temperature during antecedent periods timeframes ______ **Page 3**

***Table S4***: Intervention effects from improved on-site sanitation on log10-transformed *E. coli* counts by sample type in strata of above or below median rainfall and temperature during 2- and 7-day antecedent periods ______ **Page 4**

***Table S5***: Intervention effects from improved on-site sanitation on log10-transformed *E. coli* counts by sample type in strata of heavy rainfall and elevated temperature during 2- and 7-day antecedent periods ______ **Page 5**

***Figure S1***: Daily precipitation and monthly mean most probable number (MPN) of *E. coli* by sample type across the study period ______ **Page 6**

***Figure S2:*** Daily temperature and monthly mean most probable number (MPN) of E. coli by sample type across the study period ______ **Page 7**

***Figure S3***: Forest plot of intervention effects from improved on-site sanitation on log10-transformed *E. coli* counts by sample type in strata of heavy rainfall and elevated temperature ______ **Page 8**

**Table S1**. Number of samples collected by sample type and study round

| Round | Round 1 | Round 2 | Round 3 | Round 4 | Round 5 | Round 6 | Round 7 | Round 8 | Round 9 | Total |
|---|---|---|---|---|---|---|---|---|---|---|
| Stored water | 1013 | 641 | 592 | 571 | 599 | 581 | 582 | 574 | 587 | 5740 |
| Food | 1094 | 0 | 0 | 329 | 206 | 0 | 0 | 0 | 0 | 1629 |
| Mother hands | 0 | 720 | 705 | 684 | 682 | 668 | 662 | 643 | 633 | 5397 |
| Child hands | 1170 | 720 | 705 | 682 | 673 | 653 | 650 | 626 | 615 | 6494 |
| Source water | 1098 | 0 | 0 | 0 | 0 | 0 | 0 | 0 | 0 | 1098 |
| Soil | 1185 | 0 | 0 | 402 | 341 | 0 | 0 | 0 | 0 | 1928 |
| Ponds | 557 | 0 | 0 | 0 | 0 | 0 | 0 | 0 | 0 | 557 |
| Flies | 395 | 0 | 0 | 0 | 0 | 0 | 0 | 0 | 0 | 395 |
| **Total** | 6512 | 2081 | 2002 | 2668 | 2501 | 1902 | 1894 | 1843 | 1835 | 23238 |

**Table S2**. Percentages of samples (total n=23,238) within each weather category [a] across 2- and 7-day antecedent periods.

| | **2 Days** | **7 Days** |
|---|---|---|
| No extreme rain | 88.0% (20459) | 77.0% (17896) |
| Extreme rain | 12.0% (2779) | 23.0% (5342) |
| No extreme temp | 85.9% (19956) | 79.5% (18473) |
| Extreme temp | 14.1% (3282) | 20.5% (4765) |
| No heavy rain | 78.6% (18274) | 64.8% (15062) |
| Heavy rain | 21.4% (4964) | 35.2% (8176) |
| No elevated temp | 69.3% (16110) | 63% (14641) |
| Elevated temp | 30.7% (7128) | 37% (8597) |
| Below median rain | 49.3% (11446) | 49.0% (11379) |
| Above median rain | 50.7% (11792) | 51.0% (11859) |
| Below median temp | 52.5% (12193) | 51.1% (11867) |
| Above median temp | 47.5% (11045) | 48.9% (11371) |

[a] Extreme rain defined as ≥ 28.20 mm, extreme temperature defined as ≥ 30.21˚C, heavy defined as rain ≥ 16.44 mm, elevated temperature defined as ≥ 29.29˚C, median 2-day rolling average rain defined as ≥ 0.27 mm, median 2-day rolling average temperature defined as ≥ 27.46˚C, median 7-day rolling average rain defined as ≥ 1.10 mm, and median 7-day rolling average temperature defined as ≥27.52˚C.

**Table S3**. Improved on-site sanitation intervention effects [a] on log10-transformed *E. coli* counts by sample type in strata of extreme rainfall and temperature [b] during 2- and 7-day antecedent periods

| | **Rainfall** | | | | | | | **Temperature** | | | | | | |
|---|---|---|---|---|---|---|---|---|---|---|---|---|---|---|
| | No Extreme Rainfall | | | Extreme Rainfall | | | | No Extreme Temperature | | | Extreme Temperature | | | |
| | **Control n** | **Sanitation n** | **Δlog10 (95% CI)** | **Control n** | **Sanitation n** | **Δlog10 (95% CI)** | **Interaction P-value** | **Control n** | **Sanitation n** | **Δlog10 (95% CI)** | **Control n** | **Sanitation n** | **Δlog10 (95% CI)** | **Interaction P-value** |
| **2 Day** | | | | | | | | | | | | | | |
| **Stored Water** | 2496 | 2581 | **-0.08 (-0.15, -0.01)** | 318 | 345 | -0.05 (-0.19, 0.09) | 0.68 | 2403 | 2468 | -0.07 (-0.14, 0.00) | 411 | 458 | -0.12 (-0.29, 0.05) | 0.53 |
| **Food** | 706 | 690 | 0.04 (-0.12, 0.19) | 116 | 117 | 0.18 (-0.21, 0.56) | 0.48 | 722 | 714 | -0.12 (-0.04, 0.27) | 100 | 93 | -0.27 (-0.61, 0.08) | **0.05** |
| **Mother Hands** | 2321 | 2386 | -0.02 (-0.10, 0.05) | 335 | 355 | -0.05 ( -0.21, 0.12) | 0.75 | 2185 | 2224 | -0.01 (-0.08, 0.07) | 471 | 517 | -0.10 (-0.23, 0.03) | **0.18** |
| **Child Hands** | 2829 | 2899 | **-0.06 (-0.13, 0.00)** | 376 | 390 | -0.06 (-0.20, 0.07) | 0.99 | 2737 | 2770 | -0.04 (-0.10, 0.03) | 468 | 519 | **-0.22 (-0.36, -0.09)** | **0.01** |
| **Soil** | 834 | 810 | -0.06 (-0.18, 0.06) | 138 | 146 | -0.04 (-0.31, 0.24) | 0.86 | 853 | 847 | -0.01 (-0.13, 0.10) | 119 | 109 | **-0.40 (-0.70, -0.15)** | **0.01** |
| **Source Water** | 523 | 492 | -0.04 (-0.12, 0.04) | 40 | 43 | -0.18 (-0.49, 0.13) | 0.40 | 561 | 529 | -0.05 (-0.13, 0.03) | 2 | 6 | - | - |
| **Ponds** | 267 | 261 | -0.07 (-0.21, 0.07) | 10 | 19 | -0.40 (-0.94, 0.13) | 0.25 | 276 | 279 | - | 1 | 1 | - | - |
| **Flies** | 173 | 191 | -0.13 (-0.44, 0.18) | 20 | 11 | -0.90 (-2.30, 0.50) | 0.29 | 192 | 196 | -0.20 (-0.53, 0.13) | 1 | 6 | - | - |
| **7 Day** | | | | | | | | | | | | | | |
| **Stored Water** | 2177 | 2307 | **-0.08 (-0.16, -0.01)** | 637 | 619 | -0.03 (-0.15, 0.09) | 0.41 | 2215 | 2274 | **-0.08 (-0.16, -0.01)** | 599 | 652 | -0.05 (-0.18, 0.07) | 0.68 |
| **Food** | 566 | 576 | 0.08 (-0.10, 0.25) | 256 | 231 | 0.09 (-0.23, 0.41) | 0.93 | 673 | 661 | 0.12 (-0.04, 0.27) | 149 | 146 | -0.16 (-0.49, 0.16) | **0.12** |
| **Mother Hands** | 2057 | 2142 | -0.02 (-0.09, 0.06) | 599 | 599 | -0.05 (-0.17, 0.07) | 0.55 | 1969 | 2002 | -0.01 (-0.09, 0.08) | 687 | 739 | -0.05 (-0.17, 0.06) | 0.50 |
| **Child Hands** | 2463 | 2590 | **-0.07 (-0.14, -0.01)** | 742 | 699 | -0.02 (-0.12, 0.08) | 0.27 | 2523 | 2550 | -0.03 (-0.10, 0.04) | 682 | 739 | **-0.19 (-0.31, -0.07)** | **0.01** |
| **Soil** | 683 | 692 | -0.05 (-0.19, 0.09) | 289 | 264 | -0.08 (-0.25, 0.10) | 0.82 | 787 | 786 | 0.00 (-0.12, 0.11) | 185 | 170 | **-0.33 (-0.54, -0.12)** | **0.01** |
| **Source Water** | 428 | 430 | -0.04 (-0.13, 0.05) | 135 | 105 | -0.07 (-0.25, 0.11) | 0.76 | 561 | 529 | -0.05 (-0.13, 0.03) | 2 | 6 | - | - |
| **Ponds** | 248 | 239 | 0.06 (-0.21, 0.10) | 29 | 41 | -0.26 (-0.70, 0.18) | 0.39 | 276 | 279 | - | 1 | 1 | - | - |
| **Flies** | 136 | 162 | 0.01 (-0.32, 0.33) | 57 | 40 | **-0.91 (-1.65, -0.17)** | **0.03** | 192 | 196 | -0.20 (-0.53, 0.13) | 1 | 6 | - | - |

[a] Bolded values indicate p-value ≤ 0.05 for effect estimates and interaction p-value ≤ 0.2.
[b] Extreme rainfall defined as ≥ 28.20 mm, extreme temperature defined as ≥ 30.21˚C. We could not estimate intervention effects on source water (tubewells), ponds and flies following periods of extreme temperature due to data sparsity.

**Table S4**. Improved on-site sanitation intervention effects [a] on log10-transformed *E. coli* counts by sample type in strata of above or below median rolling average rainfall and temperature [b] during 2- and 7-day antecedent periods

| | Rainfall | | | | | | | Temperature | | | | | | |
|---|---|---|---|---|---|---|---|---|---|---|---|---|---|---|
| | Below Median Rainfall | | | Median and Above Rainfall | | | | Below Median Temperature | | | Median and Above Temperature | | | |
| | **Control n** | **Sanitation n** | **Δlog10 (95% CI)** | **Control n** | **Sanitation n** | **Δlog10 (95% CI)** | **Interaction P-value** | **Control n** | **Sanitation n** | **Δlog10 (95% CI)** | **Control n** | **Sanitation n** | **Δlog10 (95% CI)** | **Interaction P-value** |
| **2 Day** | | | | | | | | | | | | | | |
| **Stored Water** | 1408 | 1422 | -0.06 (-0.14, 0.03) | 1406 | 1504 | **-0.09 (-0.17, -0.01)** | 0.52 | 1507 | 1490 | -0.08 (-0.16, 0.01) | 1307 | 1436 | **-0.09 (-0.17, 0.00)** | 0.85 |
| **Food** | 398 | 392 | 0.18 (-0.02, 0.38) | 424 | 415 | 0.04 (-0.17, 0.24) | 0.27 | 444 | 428 | 0.06 (-0.11, 0.22) | 378 | 379 | 0.03 (-0.22, 0.27) | 0.83 |
| **Mother Hands** | 1325 | 1368 | 0.02 (-0.07, 0.11) | 1331 | 1373 | -0.06 (-0.15, 0.02) | **0.12** | 1410 | 1424 | 0.02 (-0.08, 0.12) | 1246 | 1317 | -0.06 (-0.14, 0.02) | **0.17** |
| **Child Hands** | 1601 | 1616 | -0.02 (-0.09, 0.05) | 1604 | 1673 | **-0.11 (-0.19, -0.02)** | **0.08** | 1710 | 1693 | -0.02 (-0.10, 0.05) | 1495 | 1596 | **-0.11 (-0.20, -0.03)** | **0.05** |
| **Soil** | 482 | 458 | -0.07 (-0.26, 0.12) | 490 | 498 | -0.04 (-0.18, 0.10) | 0.83 | 523 | 494 | 0.00 (-0.15, 0.14) | 449 | 462 | -0.13 (-0.30, 0.03) | 0.26 |
| **Source Water** | 277 | 241 | -0.02 (-0.13, 0.09) | 286 | 294 | -0.10 (-0.22, 0.02) | 0.35 | 311 | 265 | -0.04 (-0.13, 0.04) | 252 | 270 | -0.10 (-0.35, 0.15) | 0.71 |
| **Ponds** | 130 | 125 | -0.02 (-0.18, 0.14) | 147 | 155 | -0.20 (-0.44, 0.03) | 0.24 | 145 | 139 | -0.09 (-0.22, 0.05) | 132 | 141 | 0.03 (-0.53, 0.58) | 0.70 |
| **Flies** | 108 | 95 | 0.01 (-0.40, 0.42) | 85 | 107 | -0.41 (-0.89, 0.06) | **0.19** | 108 | 102 | -0.22 (-0.61, 0.17) | 85 | 100 | -0.13 (-0.68, 0.43) | 0.80 |
| **7 Day** | | | | | | | | | | | | | | |
| **Stored Water** | 1402 | 1414 | -0.07 (-0.15, 0.02) | 1412 | 1512 | **-0.09 (-0.17, -0.01)** | 0.67 | 1461 | 1463 | -0.07 (-0.15, 0.02) | 1353 | 1463 | **-0.08 (-0.17, 0.00)** | 0.78 |
| **Food** | 392 | 387 | 0.11 (-0.10, 0.32) | 430 | 420 | 0.00 (-0.20, 0.20) | 0.42 | 416 | 428 | 0.12 (-0.06, 0.30) | 406 | 379 | 0.02 (-0.20, 0.24) | 0.49 |
| **Mother Hands** | 1313 | 1370 | 0.02 (-0.07, 0.11) | 1343 | 1371 | -0.06 (-0.15, 0.02) | **0.11** | 1363 | 1399 | 0.00 (-0.09, 0.10) | 1293 | 1342 | -0.05 (-0.13, 0.04) | 0.37 |
| **Child Hands** | 1589 | 1613 | -0.03 (-0.10, 0.04) | 1616 | 1676 | **-0.10 (-0.17, -0.02)** | **0.11** | 1648 | 1665 | -0.04 (-0.12, 0.03) | 1557 | 1624 | **-0.09 (-0.17, 0.00)** | 0.37 |
| **Soil** | 473 | 455 | -0.05 (-0.25, 0.15) | 499 | 501 | -0.08 (-0.21, 0.06) | 0.82 | 499 | 493 | 0.03 (-0.12, 0.18) | 473 | 463 | **-0.17 (-0.31, -0.03)** | **0.05** |
| **Source Water** | 278 | 244 | -0.02 (-0.13, 0.09) | 285 | 291 | -0.10 (-0.21, 0.02) | 0.36 | 294 | 262 | -0.05 (-0.13, 0.04) | 269 | 273 | -0.08 (-0.32, 0.16) | 0.80 |
| **Ponds** | 131 | 118 | 0.00 (-0.18, 0.17) | 146 | 162 | **-0.25 (-0.48, -0.03)** | **0.10** | 136 | 138 | -0.06 (-0.20, 0.07) | 141 | 142 | -0.32 (-0.78, 0.14) | 0.30 |
| **Flies** | 96 | 104 | 0.03 (-0.39, 0.45) | 97 | 98 | -0.45 (-0.94, 0.04) | **0.15** | 102 | 100 | -0.27 (-0.67, 0.13) | 91 | 102 | 0.05 (-0.53, 0.63) | 0.40 |

[a] Bolded values indicate p-value ≤ 0.05 for effect estimates and interaction p-value ≤ 0.2.

[b] Median 2-day rolling average rain defined as ≥ 0.27 mm, median 2-day rolling temperature defined as ≥ 27.46˚C, median 7-day rolling average rain defined as ≥ 1.10 mm and median 7-day rolling average temperature defined as ≥27.52˚C.

**Table S5**. Improved on-site sanitation intervention effects [a] on log10-transformed *E. coli* counts by sample type in strata of heavy rainfall and elevated temperature [b] during 2- and 7-day antecedent periods.

| | **Rainfall** | | | | | | | **Temperature** | | | | | | |
|---|---|---|---|---|---|---|---|---|---|---|---|---|---|---|
| | No Heavy Rainfall | | | Heavy Rainfall | | | | No Elevated Temperature | | | Elevated Temperature | | | |
| | **Control n** | **Sanitation n** | **Δlog10 (95% CI)** | **Control n** | **Sanitation n** | **Δlog10 (95% CI)** | **Interaction P-value** | **Control n** | **Sanitation n** | **Δlog10 (95% CI)** | **Control n** | **Sanitation n** | **Δlog10 (95% CI)** | **Interaction P-value** |
| **2 Day** | | | | | | | | | | | | | | |
| **Stored Water** | 2214 | 2345 | **-0.08 (-0.15, -0.01)** | 600 | 581 | -0.04 (-0.15, 0.07) | 0.53 | 2027 | 2114 | -0.05 (-0.13, 0.02) | 787 | 812 | **-0.13 (-0.26, -0.01)** | 0.27 |
| **Food** | 598 | 604 | 0.03 (-0.14, 0.20) | 224 | 203 | 0.21 (-0.08, 0.50) | 0.28 | 625 | 616 | 0.16 (-0.01, 0.32) | 197 | 191 | -0.22 (-0.50, 0.06) | **0.02** |
| **Mother Hands** | 2062 | 2143 | -0.02 (-0.10, 0.05) | 594 | 598 | -0.02 (-0.14, 0.10) | 0.95 | 1777 | 1838 | -0.01 (-0.10, 0.07) | 879 | 903 | -0.05 (-0.14, 0.04) | 0.50 |
| **Child Hands** | 2518 | 2623 | -0.06 (-0.13, 0.00) | 687 | 666 | -0.07 (-0.18, 0.04) | 0.94 | 2317 | 2375 | -0.04 (-0.11, 0.03) | 888 | 914 | **-0.13 (-0.24, -0.02)** | **0.14** |
| **Soil** | 704 | 717 | -0.07 (-0.20, 0.06) | 268 | 239 | -0.04 (-0.25, 0.16) | 0.83 | 729 | 733 | 0.00 (-0.12, 0.12) | 243 | 223 | **-0.26 (-0.45, -0.06)** | **0.03** |
| **Source Water** | 467 | 463 | -0.04 (-0.12, 0.04) | 96 | 72 | -0.10 (-0.35, 0.15) | 0.64 | 0 | 41 | -0.05 (-0.13, 0.04) | 545 | 512 | -0.23 (-0.69, 0.23) | 0.44 |
| **Ponds** | 233 | 246 | -0.04 (-0.19, 0.11) | 44 | 34 | -0.13 (-0.32, 0.05) | 0.45 | 270 | 275 | -0.08 (-0.22, 0.05) | 7 | 5 | **-0.38 (-0.59, -0.17)** | **0.01** |
| **Flies** | 154 | 183 | -0.11 (-0.44, 0.22) | 39 | 19 | -0.71 (-1.56, 0.13) | **0.20** | 187 | 186 | -0.22 (-0.56, 0.12) | 6 | 16 | 0.20 (-0.63, 1.03) | 0.37 |
| **7 Day** | | | | | | | | | | | | | | |
| **Stored Water** | 1830 | 1967 | **-0.08 (-0.16, 0.00)** | 984 | 959 | -0.05 (-0.14, 0.04) | 0.58 | 1714 | 1777 | -0.07 (-0.15, 0.01) | 1100 | 1149 | -0.09 (-0.18, 0.01) | 0.79 |
| **Food** | 450 | 463 | 0.09 (-0.10, 0.28) | 372 | 344 | 0.08 (-0.15, 0.32) | 0.99 | 547 | 546 | 0.16 (-0.01, 0.32) | 275 | 261 | -0.09 (-0.36, 0.18) | **0.11** |
| **Mother Hands** | 1735 | 1809 | -0.01 (-0.09, 0.07) | 921 | 932 | -0.05 ( -0.14, 0.04) | 0.44 | 1442 | 1478 | -0.01 (-0.10, 0.08) | 1214 | 1263 | -0.04 (-0.12, 0.05) | 0.60 |
| **Child Hands** | 2084 | 2199 | -0.06 (-0.12, 0.01) | 1121 | 1090 | -0.08 (-0.17, 0.01) | 0.62 | 1958 | 1997 | -0.05 (-0.12, 0.02) | 1247 | 1292 | **-0.08 (-0.17, 0.01)** | 0.56 |
| **Soil** | 544 | 560 | -0.04 (-0.20, 0.11) | 428 | 396 | -0.08 (-0.23, 0.08) | 0.76 | 632 | 640 | 0.00 (-0.13, 0.14) | 340 | 316 | **-0.18 (-0.33, -0.03)** | **0.06** |
| **Source Water** | 369 | 377 | -0.02 (-0.12, 0.07) | 194 | 158 | -0.10 (-0.26, 0.06) | 0.42 | 521 | 493 | -0.05 (-0.14, 0.03) | 42 | 42 | -0.06 (-0.42, 0.31) | 0.98 |
| **Ponds** | 214 | 203 | -0.03 (-0.20, 0.13) | 63 | 77 | -0.22 (-0.47, 0.03) | 0.24 | 261 | 273 | -0.08 (-0.22, 0.05) | 16 | 7 | **-0.31 (-0.50, -0.13)** | **0.04** |
| **Flies** | 115 | 143 | 0.01 (-0.35, 0.37) | 78 | 59 | **-0.62 (-1.19, -0.05)** | **0.07** | 179 | 183 | -0.22 (-0.57, 0.13) | 14 | 19 | -0.04 (-0.75, 0.66) | 0.67 |

[a] Bolded values indicate p-value ≤ 0.05 for effect estimates and interaction p-value ≤ 0.2.

[b] Heavy rainfall defined as ≥ 28.20 mm and elevated temperature defined as ≥ 29.29˚C

**Figure S1**: Daily precipitation and monthly mean most probable number (MPN) of *E. coli* by sample type across the study period

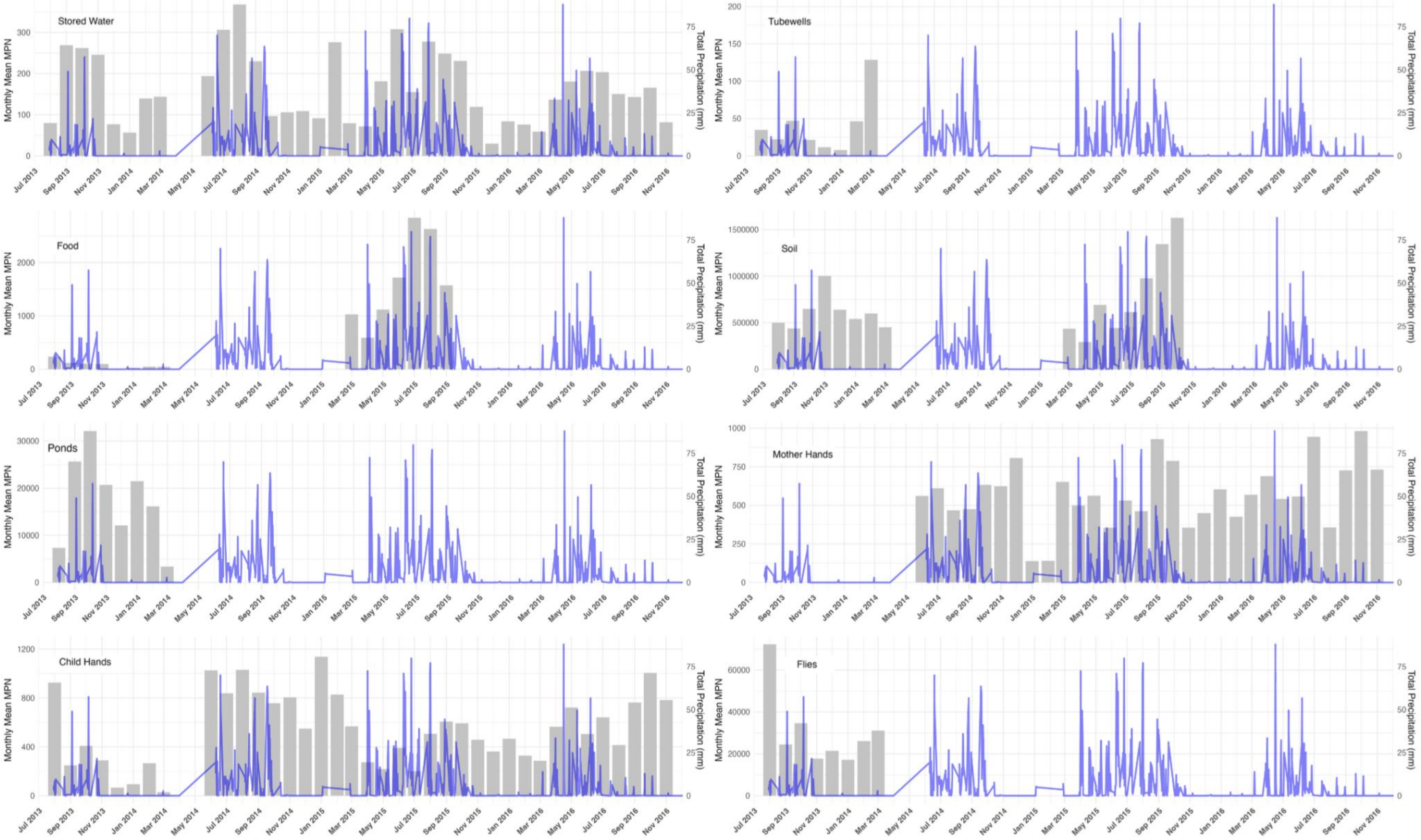

**Figure S2**: Daily temperature and monthly mean most probable number (MPN) of *E. coli* by sample type across the study period

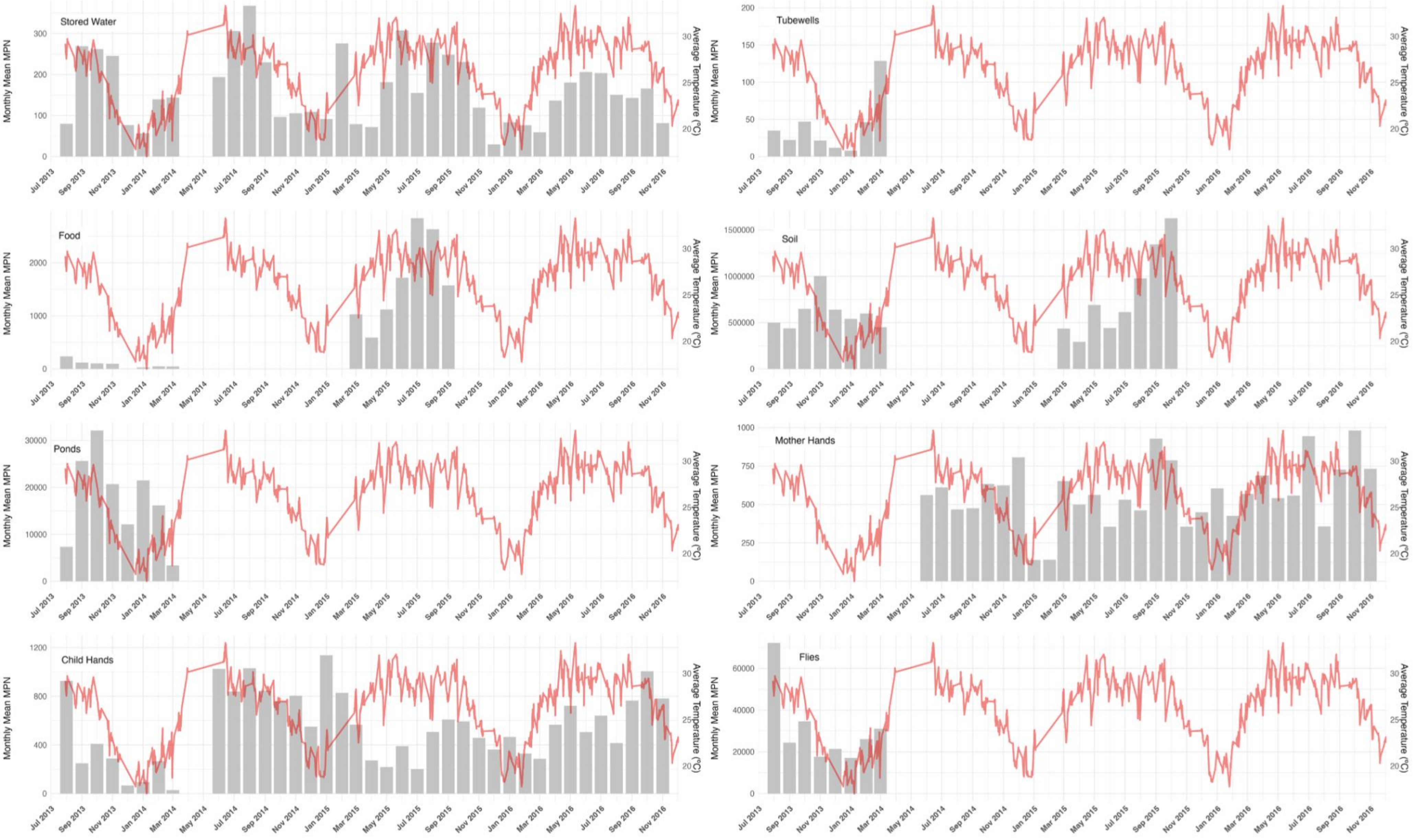

**Figure S3**. Forest plot of intervention effects from improved on-site sanitation on log10-transformed *E. coli* counts by sample type in strata of heavy rainfall and elevated temperature [a]

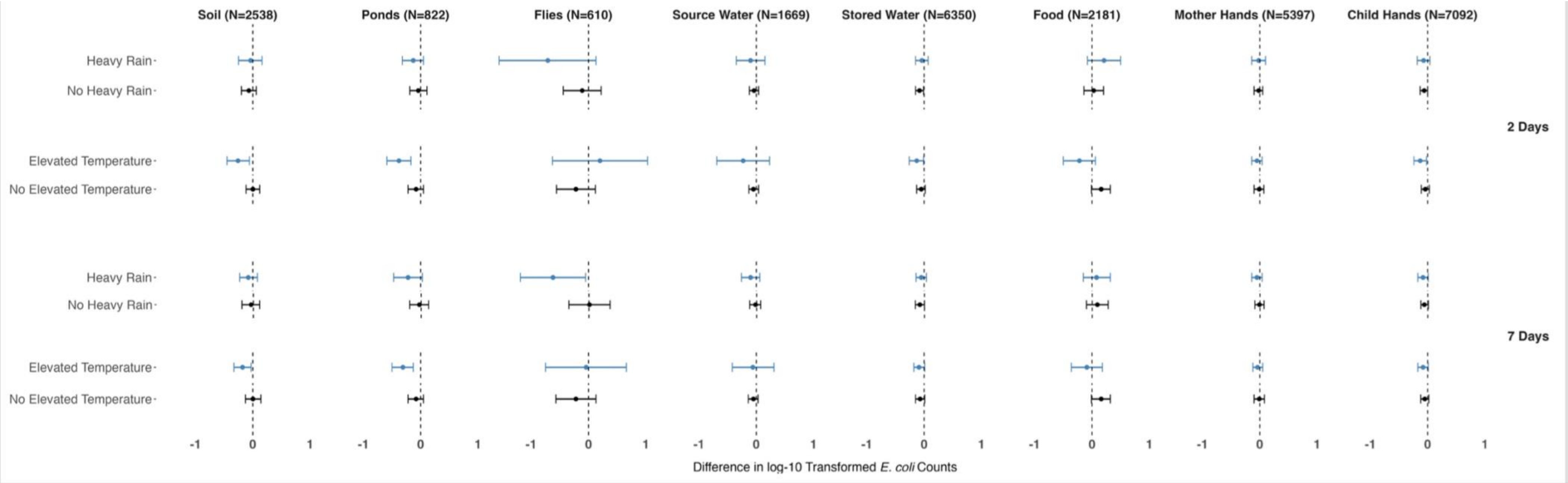


[a] Heavy rainfall defined as ≥ 16.44 mm and elevated temperature defined as ≥ 29.29˚C.
